# Boosting SARS-CoV-2 qRT-PCR detection combining pool sample strategy and mathematical modeling

**DOI:** 10.1101/2020.08.16.20167536

**Authors:** Isadora Alonso Correa, Tamires de Souza Rodrigues, Alex Queiroz, Leon de França Nascimento, Thiago Wolff, Rubens Nobumoto Akamine, Sergio Noboru Kuriyama, Luciana Jesus da Costa, Antonio Augusto Fidalgo-Neto

**Affiliations:** Instituto Senai de Inovação em Química Verde, Rio de Janeiro, 20271-030, Brazil; Instituto de Microbiologia Paulo de Góes, Departamento de Virologia, Unive+rsidade Federal do Rio de Janeiro, 21941-617, Brazil; Centro de Inovação SESI em Saúde Ocupacional, Rio de Janeiro, 20271-030, Brazil; Centro de Pesquisa Leopoldo Américo Miguez de Mello (CENPES) - Petrobrás, Rio de Janeiro, 21941-915, Brazil

## Abstract

qRT-PCR is the gold standard technique available for SARS-CoV-2 detection. However, the long test run time and costs associated with this type of molecular testing are a challenge in the actual pandemic scenario. Due to high testing demand, pooling sample strategy is an interesting approach to allow cost savings. We aim to evaluate pooling tests in experimental procedures, as well as perform *in silico* statistical modeling analysis validated with specimen samples obtained from a mass testing program of Industry Federation of the State of Rio de Janeiro (Brazil). Although the sensitivity reduction in samples pooled with 32 individuals was observed, the high-test sensitivity is maintained even when 16 and 8 samples were pooled. The *in silico* analysis showed high-cost savings in populations with positive rates lower than 15.0% according to the pool size. This data was validated with the results obtained in our mass testing program: statistical modeling predicted a cost saving of 48.0%, which in practice, was 51.5%, already considering the expenditures with pool sampling that were analyzed individually. Our data confirmed that mathematical modeling is a powerful strategy to improve the pooling approach for SARS-CoV-2 mass testing around the world while maintaining high sensitivity and robustness.

## Introduction

Since the first reported cases in Wuhan (China) in December 2019, COVID-19 (Coronavirus Disease 19) has spread around the world, and in March 2020 was classified by the World Health Organization (WHO) as a pandemic^1^. By July 2020, the number of infected people by Severe Acute Respiratory Syndrome Coronavirus 2 (SARS-CoV-2) exceeded more than 14 million people worldwide, with the accumulated COVID-19 related death over than 600,000 cases, affecting the global economies, especially those with weak health systems and social inequality^2^. Since then, the search for a pharmaceutical treatment and a vaccine against SARS-CoV-2 became the Holy Grail for research groups. However, the only effective management to control the spread of COVID-19 is contact tracing, quarantine, and social distance^3^.

Mass testing became a critical strategic approach for epidemic control, pushing the global demand for diagnosis tests exceeding the supply capacity^4^. Therefore, the pandemic information data among countries presents vast discrepancies, as the supply chain does not meet global needs. As far as other variables must be considered, one can exemplify this difference comparing the top two countries of cumulative COVID-19 cases. For example, Brazil is the top two global rank in the number of cumulative cases and deaths, behind only the USA^2^. Nevertheless, while the USA performed 150,753 diagnostic tests per million inhabitants, Brazil performed only 23,095 tests per million inhabitants, mostly in selected symptomatic and acute patients^5^.

Virus detection in human respiratory tract samples is the standard diagnosis of ongoing acute infection, which represents a reliable approach to manage virus transmission^6^. WHO recommends quantitative Reverse-Transcription Polymerase Chain Reaction (qRT-PCR) as the standard method for molecular SARS-CoV-2 detection due to its high sensibility and specificity^7^. However, the qRT-PCR cost demand could act as a limiting factor to test larger populations contributing to underreporting in the entire world. Indeed, that is the major challenge for emerging economies as diagnostic facilities, and prohibitive costs of consumables (mostly imported) is a limiting factor. Thus, extensive testing is essential, not only for transmission control but also to ensure the return of normal activities after lockdown. Pooling tests could be applied for screening large numbers of individuals during diagnosis routine, aiming to solve the experimental expenditures and supply limitations. These methodologies are based on the combination of multiple specimens in single sample analysis^8,9^. Pooling protocol has several advantages, being less expensive and time-consuming than individual testing^10^. Furthermore, this methodology is ideal for countries to become capable of performing mass testing and consequently reducing contamination levels. However, dilution of samples could affect the sensitivity of the method, as well as the amount of false-negative results.

In this work, we describe a detailed optimization of pooling test protocol to offer an economically viable approach for reliable massive diagnosis services according to the prevalence of positive cases on the evaluated population.

## Results

### • Evaluation of pooling samples with isolated SARS-CoV-2

The sensitivity test showed accurate performance when samples were pooled (Figure S1). Serial dilution curves of isolated SARS-CoV-2 showed higher sensitivity of the N1 gene, in comparison with the N2 gene, given that CT values are consistently lower for N1. For both viral targets, the detection limit was low, achieving 0.001 infectious viral particles/mL for both targets (Figure S1A), assuming that there are 10^3^ less infectious particles than total viral particles in a given viral stock, the detection limit of this test is 1 RNA copy. When initial concentrations between 100 and 10,000 infectious viral particles per mL were pooled with negative samples, a characteristic increase in CT was observed, but still within the detection limits for SARS-CoV-2 (Figure S1B-C).

### • Evaluation of pooling samples with clinical samples

Absolute CT values for all the sample pools and RNA pools tested are summarized in Tables S1 and S2, respectively. It was observed that samples with CT higher than 34 reduced the test sensitivity when 16 or 32 patients were pooled (Figure 1). Pooling strategy showed that for samples with individual CT below 29, SARS-CoV-2 detection was performed with high sensitivity, when mixing up to 32 samples into a single pool, considering the N1 gene, providing positive diagnostic according to CDC criteria^7^. However, for lower viral loads – samples with individual CT higher than 29, generally observed in patients at the beginning and the end of the infection process, poor results were found for pools consisting of 16 or 32 specimens. Detection of the N1 gene detection was possible in 86% of samples for the 1:16 pool and 57% of samples for the 1:32 pool. The sensitivity of the test also decreased for the N2 gene, with detection possible in 86% of samples for the 1:8 pool, 57% for the 1:16 pool, and 43% for the 1:32 pool.

**Figure 1.**
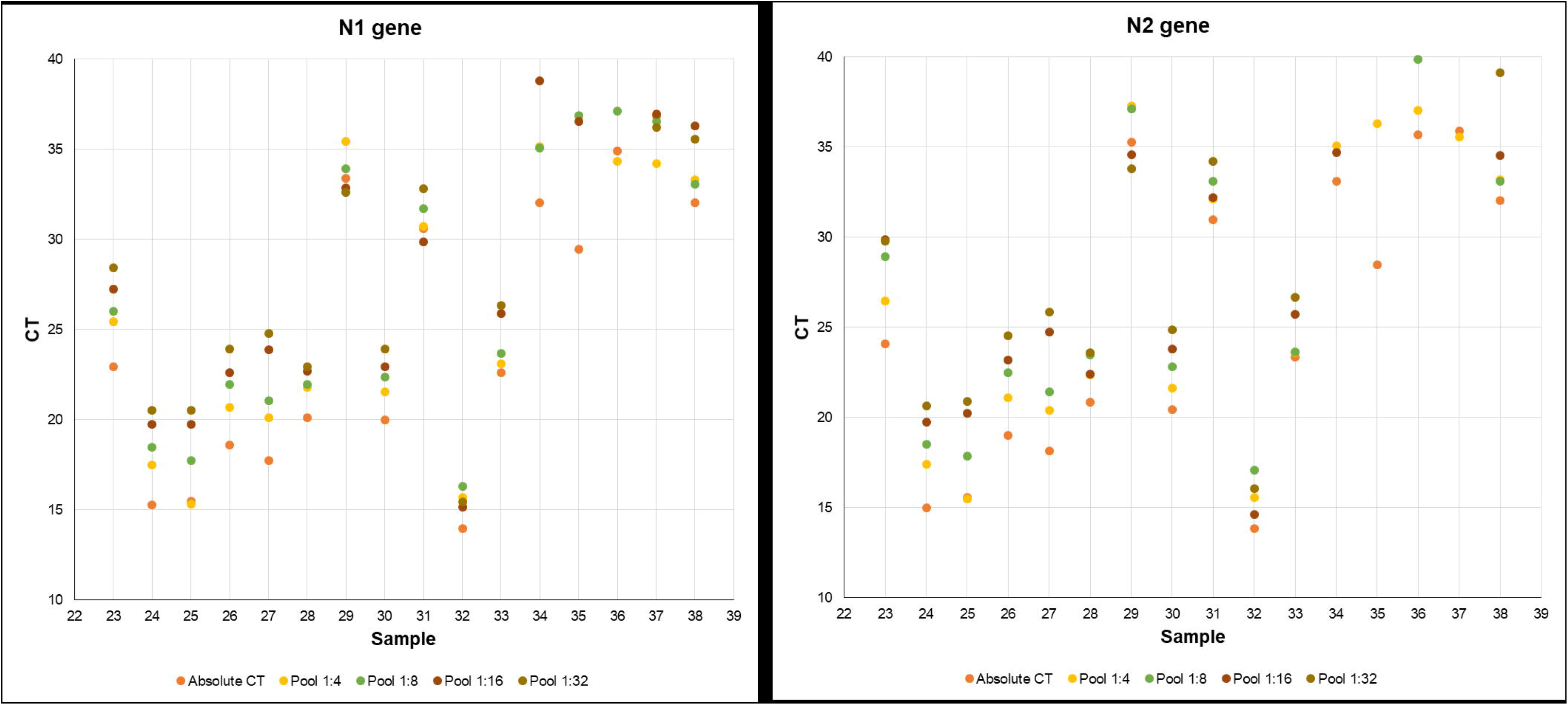
Pooling test examples of samples with distinct CT ranges. The values from all samples tested are presented in Table S1. 1:4, 1:8, 1:16 and 1:32 correspond to one positive sample pooled with other 3, 7, 15 and 32 negative samples, respectively.

PCA analysis performed with pooled samples and pooled RNA samples did not detect any group pattern among them (Figure S2; PC-1= 98% and PC-2= 1%), which means that no difference was observed between both pooling approaches. Negative and positive controls, as well as the CT curve from pools using only negative patient samples, were validated in our analyses (Figure S3).

### • In silico pooling analyses

To assess the advantages of the pooling approach, we used previous qRT-PCR results obtained in the diagnostic analyses performed with industrial workers of Rio de Janeiro state as a base to calculate the prevalence rates (%) of positive cases and to build the statistical modeling methodology. According to the in-silico methodology established, it was possible to construct a matrix evaluating the cost-savings for each pool size given positive cases prevalence. Based on this matrix, it is possible to suggest ideal pool sizes according to the prevalence rates and the cost-saving percentages on any given population (Table 1).

**Table 1.** Matrix representing pool size recommendation according to the pool size and prevalence rate of positive cases. Matrix values represent the predicted cost savings for each condition.

| Pool size | Prevalence rate of positive cases (%) |  |  |  |  |  |
| --- | --- | --- | --- | --- | --- | --- |
|  | 1 | 2.5 | 5 | 7.5 | 10 | 15 |
| 2 | 48.01 | 45.06 | 40.25 | 35.56 | 31.00 | 22.25 |
| 4 | 71.06 | 65.37 | 56.45 | 48.21 | 40.61 | 27.20 |
| 8 | 79.77 | 69.17 | 53.84 | 41.10 | 30.55 | 14.75 |
| 12 | 80.31 | 65.47 | 45.70 | 30.90 | 19.91 | 5.89 |
| 16 | 78.90 | 60.44 | 37.76 | 22.48 | 12.28 | 1.18 |
| 20 | 76.79 | 55.27 | 30.85 | 16.03 | 7.16 | -1.12 |
| 24 | 74.40 | 50.30 | 25.03 | 11.23 | 3.81 | -2.14 |
| 28 | 71.90 | 45.65 | 20.21 | 7.70 | 1.66 | -2.52 |
| 32 | 69.37 | 41.35 | 16.25 | 5.13 | 0.31 | -2.57 |

This mathematical modeling shows that populations with prevalence rates as low as 1% may reduce costs up to 80% using up to 8 or 12 specimens per pool. However, as the prevalence rate increases, the cost saving is drastically reduced in pools with a large number of samples. The need to process single analysis from the pool to identify the positive individuals increases the overall cost. Considering prevalence values equal to 5, 7.5, and 10%, the best results were observed for 1:4 pooling, with an economy of 57, 48, and 41%. As such, for prevalence rates higher than 1% but lower than 10%, pooling sizes of 4 and 8 return better cost savings in comparison to larger pool sizes. The cost modeling also demonstrates that as both pool sizes and population positive prevalence increases, the savings become marginally lower until they surpass the value of a single test, limiting the optimal cost savings to a well-defined bounded range (Figure 2).

**Figure 2.**
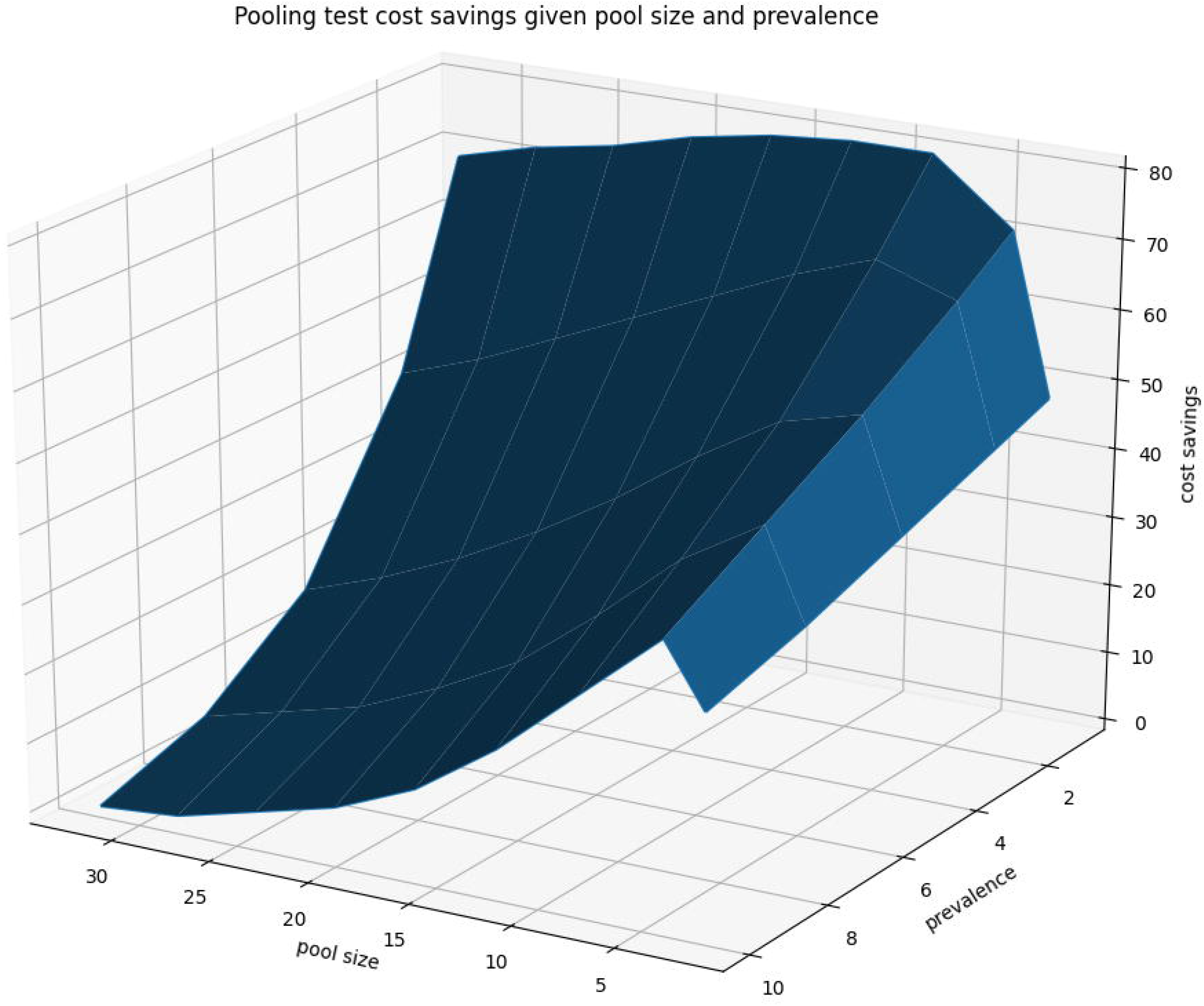
Pooling test savings given pool size and prevalence. Cost savings surface depicting optimal savings crests at pool sizes around 4 and 8. For populations with prevalence around 1%, many pool sizes are profitable, but as prevalence increases, costs savings are drastically reduced.

### • Validation of pooling strategies with clinical samples

Previous data of the COVID-19 diagnostic performed with industrial workers showed a prevalence of 7.8% positive cases in the evaluated population. According to the *in silico* analysis performed, pooling using four samples was the best choice for cost optimization (Table 1). To assess the precision of in silico model, the mass testing program for industrial workers of Rio de Janeiro State was tested using the pooling strategy. A total of 6,096 samples were processed at the SESI Innovation Center for Occupational Health, constructing 1,524 pools (Table 2). From those pools, 365 were positive (24.0%), which implicates in more 1,460 qRT-PCR tests to identify the positives samples. Overall, an economy in 51.1% was observed using this strategy (Table 2). This result agrees with the statistical modeling prediction performed (Table 1).

**Table 2.**
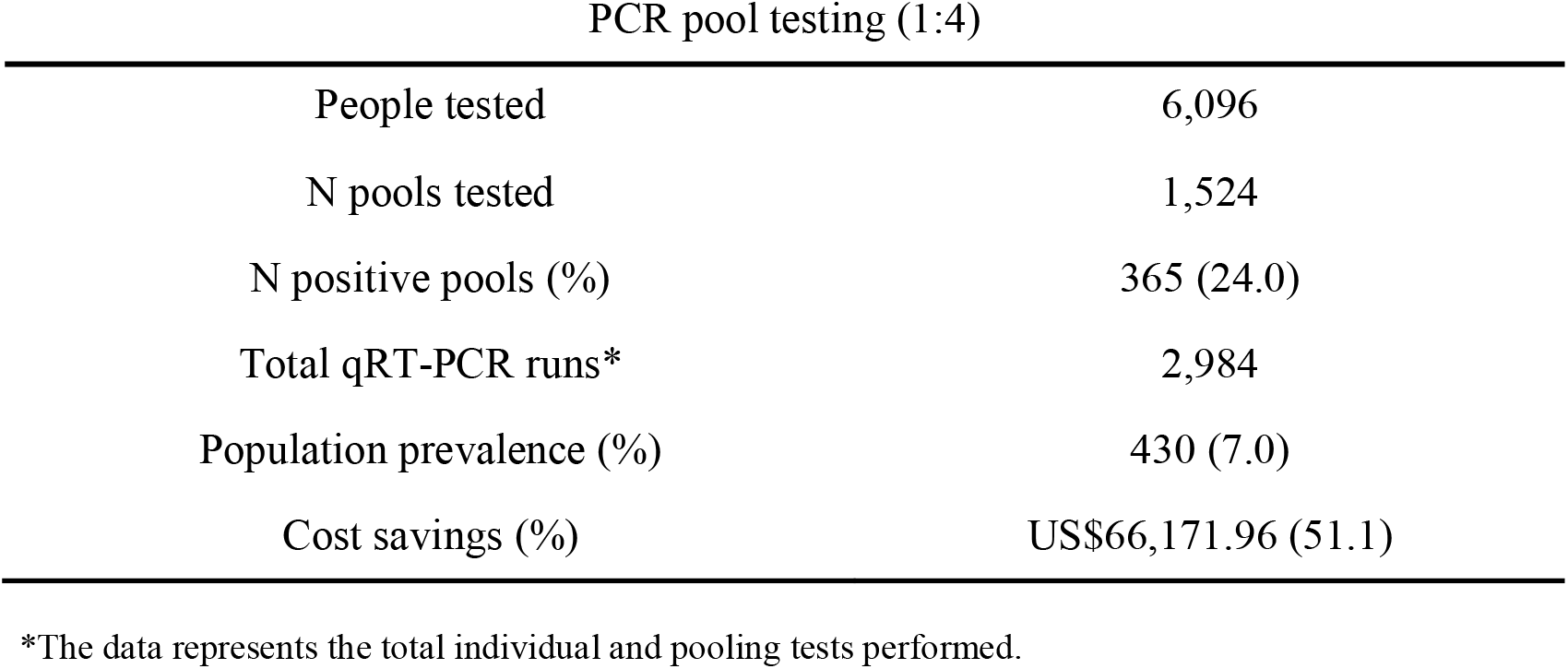
The pooling strategy applied to thr testing program for industrial workers of Rio de Janeiro State.

## Discussion

Diagnostic of COVID-19 is a crucial factor for not only control the spread of pandemic but also for evaluating the process of restore activities and monitor possible new waves of infection. However, limited access to reagents and consumables delay testing and affect these actions. This problem is verified when we analyze the positive results of people tested, known as the positive rate of different countries. While some countries like Australia, South Korea, and Uruguay present a positive rate of less than 1%, others show rates bigger than 10% (Sweden, 12.1% and South Africa, 21.9%), even reaching 55.8% like Mexico meaning that these countries are not testing enough^11^. For Brazil, this data is not available, but until June, 1.48 million tests were performed, a rate of 6.96 tests per thousand people^11^. Although test numbers are increasing, this rate is still insufficient, especially considering that a percentage of these tests are serological, which are not ideal for the diagnostic of active infections. Mass testing needs to be implemented to detect the transmission of SARS-CoV-2 in the community and to provide multi-time point surveillance.

Dorfman designed the strategy of pooling samples in 1940 to screening for syphilis infection in large populations of soldiers^12^. Since then, pooling is widely used for the detection of other pathogens for diagnostic purposes and even to study the prevalence of these agents in a defined population^9,13,14^. In a pandemic situation, as for COVID-19, large-scale diagnostic testing is strongly recommended to define the prevalence rates in a region and to control the spread of the infection. Moreover, qRT-PCR-based molecular testing is the only test indicated to ascertain correctly SARS-CoV-2 infected individuals with transmission potential. However, this aim is hard to reach because of the vast numbers of tests required globally in a short period, resulting in a scenario of reagents, equipment, and consumables deficit and an overload for diagnostic laboratories around the world. This way, pooling samples may be an interesting approach to overcome these limitations and provide access to mass testing.

One of the main concerns of pooling is the size of assembled pools that should be evaluated according to the prevalence of the pathogen in the study population. Some studies have demonstrated that for diseases with low population prevalence, this approach has the most potential for enabling mass testing at low costs, including for SARS-CoV-2 detection^10,15^. Besides, it was seen a great potential of pooling for repeat testing of the same population on a consecutive period, which could be an efficient strategy for disease control^16^. However, the prediction of ideal pool size (Table 1) requires a discerning in silico analysis. Otherwise, the cost-saving will not be reached. Mathematically, applying the Dorfman’s approaches may incur savings as high as 90% within populations with a prevalence close to 1%. Our study adopted the statistical modeling approach and validated the data with pooling biological samples for COVID-19 diagnostic, confirming that the pool size must be selected according to the prevalence rate of positive cases in the population (Figure 2). Besides, this combined analysis is essential to allow the optimization of limited resources and to apply mass testing enabling the management and reduction of underreporting, observed globally, but especially in large and developing countries^17,18^. For SARS-CoV-2 detection, other published studies performed pooling test validations based on mathematical models. Abdalhamid and colleagues (2020) used a web-based application to calculate pool size and reach similar results to ours with a recommended pool size of five samples for a 5% prevalence rate and an economy of 57% in tests. Other studies propose pooling RNA using two up to 64 samples^19,20^.

Although Yelin and coworkers (2020) have demonstrated the possibility of making pools using RNA or swab samples with the same test quality, the authors used a limited number of samples^20^. Here, our results showed that pooling RNA or nasopharyngeal swab samples has the same efficiency for SARS-CoV-2 diagnostic, which chooses pooling nasopharyngeal swab samples a way to overcome the RNA extraction bottleneck. Another critical factor is sample dilution that might be responsible for inconclusive or even false-negative results, depending on the CT range of the analyzed samples. In this work, we thoroughly tested different concentrations of viral stock preparations and clinical samples in pools up to 32 samples. We demonstrated that even at the concentrations close to the qRT-PCR detection limit (pools up to 8 samples) were still positive. Larger pools could also lead to problems related to samples with low viral load, whose detection could be missed, and the logistic of assembly and deconvolution of these pools needs to be done carefully to avoid cross-contamination. A study proposes the deconvolution of pools divided into stages according to prevalence rate could optimize the process and increase the samples pooled^21^. Nevertheless, adding more steps to the diagnostic chain could delay the final diagnostic and spend more resources.

Our study associates *in silico* analyses and test validation to ensure an overall and safe methodology to be widely used, and one that will reduce false and inconclusive diagnostic results, and save costs. We performed the implementation of pool methodology in the COVID-19 mass testing program of the Industry Federation of the State of Rio de Janeiro. We observed substantial gains by reducing the qRT-PCR run time and the use of reagents and general consumables (Table 2). The statistical model predicted a cost saving of 48% (Table 1) for the pooling of four nasopharyngeal samples in a population of 7.5% of the prevalence rate. Indeed, we are observing an economy of 51.1% in our test routine. This cost-saving already considers all the subsequent re-testing for the diagnosis of individual samples and the false-negative rate. In our analysis, the false-positive rate was 15.28%, meaning that 57 of 373 pools that presented a CT curve were negative when samples were analyzed individually. This rate is higher than previously reported by other authors^22,23^ but could be explained by the higher number of tests performed in our routine. Besides, we established a stringent parameter for the pooling test interpretation, for which any positive signal regardless of the CT and the fluorescence signal was considered positive, even those that would be considered inconclusive in individual tests. Therefore, excluding results with CTs above 34 in the pooling test, the rate drops to 5.19%, it is within the range of other studies^22,23^. Given that the test cost of COVID-19 diagnostic in-house PCR used in our company is around US$22.20 per individual sample, pooling samples by four reduce the experimental cost to US$10.85 per test. This reduction of approximately twice was also observed in another study for a prevalence of 5%^24^.

In summary, our study demonstrates that the pooling strategy, associated with a robust *in silico* analysis, can boost testing efficiency and save resources in populations with a low prevalence of positive cases of COVID-19, in addition to reducing costs. This approach helps to implement mass testing and detect the transmission of SARS-CoV-2 in the community, as well as the epidemic multi-time point surveillance, supporting actions both to control the spread of the infection and to define adjustments in social isolation measures.

## Methods

### • Samples

Clinical samples were collected from the mass testing program of Industry Federation of the State of Rio de Janeiro (Brazil), from industry workers, between April and May 2020. The nasopharyngeal swabs were conditioned in DMEM medium (Thermofisher), and 1.5 mL of each sample were individually stored at -80°C in cryotubes until further use. For the pooling validation, we used leftovers from routine testing samples, and no personal, clinical, and demographic data from individuals were accessed, therefore the institutional ethics committee (CEP/HUCFF/FM/UFRJ) waived the informed consent term for this study. All methods and experimental protocols were approved and carried out in accordance with guidelines and regulations from the institutional committee.

### • Sensitivity tests

SARS-CoV-2 was previously isolated from culture cells and titrated in the Molecular Virology Laboratory, Federal University of Rio de Janeiro (Rio de Janeiro – Brazil). The total amount of 10^6^ viral infectious particles per mL were serially diluted (10^-1^, 10^-2^, 10^-3^, 10^-4^, 10^-5^, 10^-6^, 10^-8^, 10^-9^ and 10^-10^ infectious viral particles/mL) to detect the maximum sensitivity of the test during pooling analyses. Furthermore, to determine the sensitivity of different pool sizes (4, 8, and 16 samples each), titers of isolated virus (curve of 100, 1,000, and 10,000 viral infectious particles/mL) were pooled with nasopharyngeal swabs previously diagnosed as negatives for COVID-19.

### • Pooling assembly strategies

Here, it was performed a comparative analysis between the nasopharyngeal sample and RNA pooling approaches to identify the best method to construct the pools.

#### Nasopharyngeal Sample Pooling

To evaluate the effect of pooling samples on the qRT-PCR sensitivity, Cycle Threshold (CT) values of pools were compared with CT values of the individual test at the same run. A total of 38 patients previously described as positive with distinct CT values were tested in the pooling test using frozen aliquots of nasopharyngeal swabs previously detected as positive or negative in the diagnostic tests. Each pool was prepared with an equal volume of one positive and others *n* negative patients, varying according to the pool size. Combinations of 4, 8, 16, or 32 specimens were tested. The final volume of 300 μL was used to perform RNA extraction.

#### RNA Pooling

For RNA pools, we selected 24 RNA samples individually extracted from nasopharyngeal swabs before the pool’s construction. Pooling was performed from RNA samples, combining one SARS-CoV-2 positive with another *n* negative RNAs, ranging according to the pool size. As well as sample pooling, we tested pools of 4, 8, 16, and 32 RNA sample combinations.

### • RNA extraction

RNA extraction was performed mechanically by Maxwell® RSC (Promega) using a sample volume of 300 μL. Total RNA was obtained by the Maxwell® RSC Viral Total Nucleic Acid Purification Kit (catalog number: AS1330 - Promega) according to the manufactured protocol. Non-pooled samples and negative control also had RNA extracted.

### • cDNA synthesis and real-time PCR

qRT-PCR reactions were performed in one step with Brilliant III Ultra-Fast qRT-PCR Master Mix (catalog number: 600884 - Agilent). Each reaction containing: 10μL of 2X QRT-PCR Master Mix; 1.5 μL prime time; 0.2μL of 100mM DTT; 0.3 of reference dye (dilute 1:500); 1.0μL of RT/RNase block; 2.0μL of nuclease-free water and 5μL of RNA. Reactions were cycled at QuantStudio 5 (Thermo Scientific) according to the following program: 50°C for 10 minutes; 95°C for 3 minutes; 45 cycles of 95°C for 3 seconds and 53°C for 30 seconds.

### • Primers and probes

Primers and probes for viral gene N (N1 and N2) were synthesized as primetime chemistry by Integrated DNA Technologies (IDT) according to sequences from the Center for Disease Control and Prevention (CDC-US). Human RNase P (hRNase P) was used as human specimen control.

### • Statistical analysis

Principal Component Analysis (PCA) was performed by using Unscrambler Software (version 10.1; CAMO AS; Trondheim, Norway). Grouping pattern was investigated within samples S23-S38, considering as variables the pool type (sample pooling or RNA pooling) and the pool size (4, 8, 16, or 32 samples).

### • In silico experimental design using pooling methodology

Mathematical modeling of the pooling strategy was performed based on pool size and prevalence of positive cases in a given population. Each pooled test was modeled as a Bernoulli trial and with the probability of a positive test *p* based on the results obtained from the clinical nasopharyngeal swab samples collected from industrial workers. The standard case was no positive cases within a pooled test, given by:

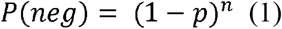

where P*(neg)* gives the probability (*p*) of a given pooled sample with size (*n*) having no positives based on the exponential distribution.

To estimate the cost savings related to a pooling strategy, the model below (2)

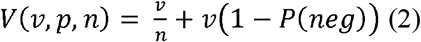

was developed to estimate the reduced test value using the pooling strategy, where *v* represents the costs for a single test, *p* is the probability of a positive test, *n* is the selected pool size, and the term *v*(1 - P(*neg*)) gives the cost of redoing tests given P(*neg*).

The cost savings are given by the ratio between the reduced test value and the single test value subtracted from the whole.

### • Pooling methodology application in clinical samples

To validate the in silico experimental design developed in this work, as well as increase the testing capacity performed by SESI Innovation Center for Occupational Health, clinical samples collected individually, as described above, were pooled using four specimens. A total of 6,096 samples were tested for SARS-CoV-2 using the RT-PCR processed as 1,524 pools (1:4).

### • Pooling test and individual test interpretation samples

In our routine, we established the CDC criteria for individual tests^7^. However, for the pooling strategy, detection of any fluorescence signal in a pool, regardless of the CT or fluorescence value, was considered positive, and the samples in the specific pool were analyzed individually.

## Data Availability

The authors confirm that the data supporting the findings of this study are available within the article [and/or] its supplementary materials.

## Acknowledgements

We thank Mr. Ronaldo Rocha for technical assistance.

## Author contributions

IAC, TSR, AQ, SNK, LJC and AAF conceived and designed the experiments. IAC and TSR carried out the RT-qPCR assays and analyzed the data. LFN made the mathematical modeling. TW performed the graphical design of figures and tables. IAC, TSR, LFN, AQ, LJC and SNK wrote the manuscript. RNA, SNK, LJC, and AAF provided intellectual input and revised the manuscript. All authors read and approved the final manuscript.

## Additional Information

### Funding

Grants from Conselho Nacional de Pesquisa (CNPq), and Carlos Chagas Filho Foundation for Research Support of Rio de Janeiro State (FAPERJ), Brazil. ISI QV and CIS HO are supported by the Industry Federation of Rio de Janeiro FIRJAN. This work is part of the CENPES/Petrobras’ Scientific Structure of Response to the COVID 19 pandemic sponsored by CENPES/PETROBRAS.

### Competing interests

The authors declare no competing financial interests.

## Notes

### Competing Interest Statement

The authors have declared no competing interest.

### Author Declarations

The institutional ethics committee (CEP/HUCFF/FM/UFRJ) oversighted the the study and waived the informed consent term.

